# Evaluating the Adoption of Voice Recognition Technology for Real-Time Dictation in a Rural Healthcare System: A Retrospective Analysis of Dragon Medical One

**DOI:** 10.1101/2022.07.22.22277923

**Authors:** Adedayo A. Onitilo, Abdul R. Shour, David S. Puthoff, Yusuf Tanimu, Adedayo Joseph, Michael T Sheehan

## Abstract

**Background:** In 2013, Marshfield Clinic Health System (MCHS) implemented the Dragon Medical One (DMO) system provided by Nuance Management Center (NMC) for Real-Time Dictation (RTD), embracing the idea of streamlined clinic workflow, reduced dictation hours, and improved documentation legibility. Since then, MCHS has observed a trend of reduced time in documentation, however, the target goal of 100% adoption of voice recognition (VR)-based RTD has not been met.

**Objective:** To evaluate the uptake/adoption of VR technology for RTD in MCHS, between 2018-2020.

**Methods:** DMO data for 1,373 MCHS providers from 2018-2020 were analyzed. The study outcome was VR uptake, defined as the number of hours each provider used VR technology to dictate patient information, used as continuous, and classified as no/yes. Covariates included sex, age, US-trained/international medical graduates, trend, specialty, and facility. Descriptive statistics and unadjusted and adjusted logistic regression analyses were performed. Stata/SE.version.17 was used for analyses. P-values less than/equal to 0.05 were considered statistically significant.

**Results:** Of the 1,373 MCHS providers, the mean (SD) age was 48.3 (12.4) years. VR uptake was higher than no uptake (72.0% vs. 28.0%). In both unadjusted and adjusted analyses, VR uptake was 4.3 times and 7.7 times higher in 2019-2020 compared to 2018, respectively (OR:4.30,95%CI:2.44-7.46 and AOR:7.74,95%CI:2.51-23.86). VR uptake was 0.5 and 0.6 times lower among US-trained physicians compared to internationally-trained physicians (OR:0.53,95%CI:0.37-0.76 and AOR:0.58,95%CI:0.35-0.97). Uptake was 0.2 times lower among physicians aged 60/above than physicians aged 29/less (OR:0.20,95%CI:0.10-0.59, and AOR:0.17,95%CI:0.27-1.06).

**Conclusion:** Since 2018, VR adoption has increased significantly across MCHS. However, it was lower among US-trained physicians than among internationally-trained physicians (although internationally physicians were in minority) and lower among more senior physicians than among younger physicians. These findings provide critical information about VR trends, physician factors, and which providers could benefit from additional training to increase VR adoption in healthcare systems.

## Introduction

Marshfield Clinic Health System (MCHS) was founded in 1916 by six physicians and has grown to become Wisconsin’s largest private group medical practice, with over 50 locations and over 300,000 patients in northern, central, and western Wisconsin. In 2013, MCHS implemented the Dragon Medical One (DMO) system provided by Nuance Management Center (NMC) for Real-Time Dictation (RTD), embracing the idea of streamlined clinic workflow, reduced dictation hours, and improved documentation legibility (1). Since then, MCHS’s information technology department has tracked the minutes of time spent on manual patient documentation by either physicians or allied healthcare staff on a weekly basis. While observing a trend of reduced time in documentation, the target goal of 100 percent adoption of voice recognition (VR)-based RTD has not been met. At the end of August 2019, 30 high-end users out of more than 250 healthcare providers continued to manually document patient care information. At MCHS, achieving 100 percent VR usability remains a challenge, and the accuracy of RTD remains a source of concern.

MCHS providers who document patient information in electronic health records (EHR) come from a variety of backgrounds and play many roles, including physicians, nurses, physician assistants, surgeons, social workers, and others. Individuals’ experience, and acceptance of VR may differ (2–4), resulting in varying training requirements. The main challenge that MCHS faced was completing the institute-wide implementation of VR technology among diverse providers working various shifts at sporadic local clinics and centers. Furthermore, our rural populations are served by a relatively high proportion of overseas-trained healthcare providers. Foreign-trained or international-trained medical graduates contribute significantly to filling medical gaps in US communities (5–7). Although healthcare providers in rural communities provide excellent care (8), whether they have received foreign-trained education influences their adoption of new technology. As a result, deciding how much and what kind of training was best for MCHS providers at dispersed worksite locations required careful consideration.

To familiarize providers with the new technology and provide proper training, MCHS distributed End-User Guides and Quick Reference Guides via email and posts to the internal website, and strategically provided three types of training at all sites: large group, small group, and one-on-one. The large group trainings were held at the Marshfield, Chippewa, Eau Claire, Minocqua, Rice Lake, and Wausau centers. Six large group trainings were conducted at Marshfield Center in 2018, and fifteen large group trainings were conducted at other centers in February and March of 2019. Trainers visited service locations to provide small group and one-on-one training to those who were unable to attend the large group trainings. Approximately 1,000 providers participated in small group and one-on-one trainings from August 2018 to June 2019. Approximately 30 completed their training at the end of 2018; approximately 600 providers completed the training between April and May of 2019. To ensure information consistency, all trainers were issued a Training Guide. Despite these efforts, some service providers continue to rely on traditional transcription services. It was unclear how far all efforts have advanced providers’ adoption of VR technology or their use of RTD as the primary measure of documentation.

Hence, our study aimed to systematically examine the overall adoption of the new technology by examining trends and user and institutional factors that influenced VR uptake at MCHS between 2018 and 2020. This study, which considers MCHS’s future implementation of the VR system, will have important implications for other health services in the country. With important information about VR uptake trends, physician factors, and which providers could benefit from additional training to increase VR adoption, promote streamlined clinic workflow, reduce dictation hours, and improve document legibility in healthcare systems across the United States.

## Materials and Methods

### Data sources, and study measures

Data were analyzed from the DMO Analytics system, which is a secure web-based analytics portal accessible via NMC that includes a suite of self-service reporting tools with data visualizations that can provide answers to key healthcare questions (9). Using a 1,373 (N) MCHS providers, this study took a retrospective approach to evaluate VR uptake data during 2018-2020. All healthcare providers from institute-wide departments, various locations, and staff types who worked between 2018 and 2020 were eligible, and this study was approved by Marshfield Clinic Institutional Review Board (IRB-20-718).

The overall study outcome/dependent variable was VR uptake, defined as the number of hours each provider used VR technology and dictated patient information. VR uptake was used as a continuous variable and classified as either no VR uptake or uptake. Individual user, institutional characteristics and VR user experience variables were used as covariates.

Individual user characteristics included sex (categorized as female and male), age used as continuous (categorized as 29 years/less, 30-39 years, 40-49 years, 50-59 years and 60 years/above), US-trained/international medical graduates (coded as international medical doctor and US-trained MD), physician/allied health, surgical/non-surgical, year started working at MCHS (coded as 1978-1988, 1989-1998, 1999-2009 and 2010-2020), still working (no/yes), historical trend (2018, 2019 and 2020), and years of practice experience from any institution including MCHS used as continuous (and categorized as 10 years/less, 11-20 years and 21 years/above).

Institutional factors were specialty/department (categorized as pediatric care and adult care specialties) and primary facilities including the Marshfield Medical Center and other MCHS facilities such as Ascension Our Lady of Victory Hospital, Ascension St. Clare’s Hospital, Ascension St. Mary’s Hospital, Ascension St. Mary’s at Crandon, Ascension St. Mary’s at Rhinelander, Bloomer Center, Cadott Center, Chetek Center, Chippewa Falls Center, Clairemont Center, Colby-Abbotsford Center, Cornell Center, Cumberland Center, Eagle River Center, Eau Claire Center, Flambeau Hospital, Greenwood Center, Hayward Center, Howard Young Medical Center, Ladysmith Center, Lake Hallie Center, Lakewoods Center, MMG-SP Illinois, MMG-Weston, Marshfield Alcohol and Drug Recovery Center, Marshfield Medical Center - Eau Claire, Marshfield Medical Center – Ladysmith, Marshfield Medical Center – Minocqua, Marshfield Medical Center – Neillsville, Marshfield Medical Center - Rice Lake, Marshfield Plaza Therapy Center, Menomonie Center, Mercer Center, Merrill Center, Minocqua Alcohol and Drug Recovery Center, Minocqua Center, Mosinee Center, Neillsville Center, Oakwood Center, Park Falls Center, Phillips Center, Rhinelander Center, Riverview Center, St. Michael’s Hospital, Stettin Center, Stevens Point Center, Stratford Center, Wausau Center, Weston Center, Wisconsin Rapids Center, Wittenberg Center, and Woodruff Center.

Voice recognition user experience variables included lines per hour of clinical documentation generated by provider, dictation hours, total lines generated, auto text number, voice commands number and signal to noise ratio (9) (all used as continuous variables). Lines/hour were defined as 65 characters per line (including spaces), and measured how many lines of clinical documentation a provider generated in a one-hour period while using VR (on average) (9). Numbers of Auto-text Commands were defined as the number of auto-text instances a provider has while dictating with VR. Numbers of Voice Commands were defined as the number of times a provider used voice commands while dictating with VR. The Signal-to-Noise Ratio was defined as the ratio of input volume to background noise value, indicating the audio quality for RTD. Greater than 10 decibels is considered acceptable (9).

### Statistical analysis

We used a five-step process to analyze our data. First, we ran descriptive statistics on all study measures, such as counts/frequency and percentages for categorical variables and means and Standard Deviations (SD) or medians for continuous variables. Second, a bivariate analysis was carried out, with chi-square tests used to compare all study variables by study outcome (VR uptake). Third, a multivariate logistic regression analysis was used to investigate the relationship between study outcome and covariates (individual user, and institutional characteristics) across MCHS. Fourth, an unadjusted analysis of user and institutional factors influencing VR uptake among MCHS physicians was conducted. Fifth, an adjusted analysis of user and institutional factors influencing VR uptake among MCHS physicians was performed. Finally, we provided percentages, p-values, unadjusted and adjusted odds ratios (AOR), and 95% confidence intervals (CIs). Stata/SE.version.17 (10) was used for all statistical analyses, and p-values of 0.05 or less were considered statistically significant.

## Results

Table 1 depicts the user and institutional characteristics of MCHS providers from 2018 to 2020. The 1,373 (N) MCHS providers had a mean (SD) age of 48.3 (12.4) years. VR uptake was higher than no VR uptake (72.0% vs. 28.0%) among the 1,373 MCHS providers. In 2018, 79.4% of MCHS providers were included in our study, compared to 17.1% in 2019 and 2020 (3.5%).

**Table 1:** User and Institutional Characteristics of MCHS Providers, 2018-2020, N=1,373.

| Study Measures | n | % |
| --- | --- | --- |
| **Individual User characteristics:** |  |  |
| Sex |  |  |
| Female | 686 | 49.96 |
| Male | 687 | 50.04 |
| Age*, n, min-max, mean (SD), median | 1310 | 22-83, 48.26 (12.40), 48 |
| Age categories<sup>a</sup> |  |  |
| 29 years/less | 68 | 6.68 |
| 30-39 years | 41 | 4.03 |
| 40-49 years | 285 | 28.00 |
| 50-59 years | 336 | 33.01 |
| 60 years/above | 288 | 28.29 |
| US-trained/international medical graduates |  |  |
| International MD | 269 | 21.42 |
| US-trained MD | 987 | 78.58 |
| Physician/Allied Health<sup>a</sup> |  |  |
| Medical Doctor (MD)/physician | 554 | 40.47 |
| Non-physician/Allied Health | 815 | 59.53 |
| Surgical/non-surgical |  |  |
| Non-Surgical | 1,275 | 92.86 |
| Surgical | 98 | 7.14 |
| Year started working at MCHS categories<sup>a</sup> |  |  |
| 1978-1988 | 49 | 3.58 |
| 1989-1998 | 156 | 11.39 |
| 1999-2009 | 326 | 23.80 |
| 2010-2020 | 839 | 61.24 |
| Still working |  |  |
| No | 259 | 18.86 |
| Yes | 1,114 | 81.14 |
| Historical trend (Months converted to years) |  |  |
| 2018 | 1,090 | 79.39 |
| 2019 | 235 | 17.12 |
| 2020 | 48 | 3.50 |
| Years of practice*, min-max, mean (SD), median | 1370 | 0-46, 10.64 (9.61), 7 |
| Years of practice categories<sup>a</sup> |  |  |
| 10 years/less | 821 | 59.93 |
| 11-20 years | 315 | 22.99 |
| 21 years/above | 234 | 17.08 |
| **Institutional characteristics:** |  |  |
| Specialty/Department<sup>a</sup> |  |  |
| Pediatric Care Specialties | 127 | 9.41 |
| Adult Care Specialties | 1,223 | 90.59 |
| Primary Facility<sup>a</sup> |  |  |
| Marshfield Medical Center | 601 | 45.43 |
| Others (e.g., Marshfield Medical Center-Eau Claire, Marshfield Medical Center-Rice Lake, Wausau Center, Weston Center, etc.) | 722 | 54.57 |
| **VR User Experience** |  |  |
| VR uptake hours., min-max, mean (SD), median | 1373 | 0-17.3, 2.16 (2.83), 1 |
| VR uptake categories |  |  |
| No VR hours | 385 | 28.04 |
| VR uptake hour | 988 | 71.96 |
| Lines/hour of clinical documentation generated<sup>a</sup>, min-max, mean (SD), median | 1137 | 0 -1878.73, 428.61 (187.46), 426.86 |
| Dictation Hours, min-max, mean (SD), median | 1373 | 0-29.30, 1.83 (3.77), 0 |
| Total lines<sup>a</sup>, min-max, mean (SD), median | 1137 | 0-13409.1, 1488.88 (1800.95), 876.10 |
| Auto Text Number<sup>a</sup>, min-max, mean (SD), median | 1137 | 0-899, 9.52 (50.49), 0 |
| Voice Commands number<sup>a</sup>, min-max, mean (SD), median | 1137 | 0-2760.5, 37.31 (128.75), 3.5 |
| Signa Noise ratio<sup>a</sup>, min-max, mean (SD), median | 1137 | 0-34.24, 15.31 (6.077), 15.46 |
| <sup>a</sup>Frequency may not add up to the total sample due to missing observations |  |  |

There were more physicians compared to non-physicians/allied health professionals (59.5% vs. 40.5%), and MDs/physicians trained in the United States than international physicians (78.6% vs. 21.4%). A total of 1878.7 lines of clinical documentation were generated per hour, with a mean (SD) of 428.6 (187.5). There were 29.3 dictation hours total, with a mean (SD) of 1.83 (3.8) and a median of 0. A total of 899 auto text messages were generated, with a mean (SD) of 9.5 (50.5) and a median of 0. There were 2760.5 voice commands generated in total, with a mean (SD) of 37.3 (128.8) and a median of 3.5. A total of 34.2 lines of Signa Noise ratio were generated, with a mean (SD) of 15.3. (6.1).

Table 2 presents a bivariate analysis of MCHS providers’ VR uptake by user and institutional characteristics between 2018 and 2020. VR uptake decreased significantly in 2019-2020 compared to 2018 (25.9% vs. 74.1%; p<0.001). Other characteristics were associated with VR uptake including sex (p=0.003), age (p<0.001), the year provider started working at MCHS (p<0.001), US-trained/international medical graduates (p=0.002), whether the provider is still working (p<0.001), years of practice experience (p<0.001), and location of the primary facility (p<0.001).

**Table 2:**
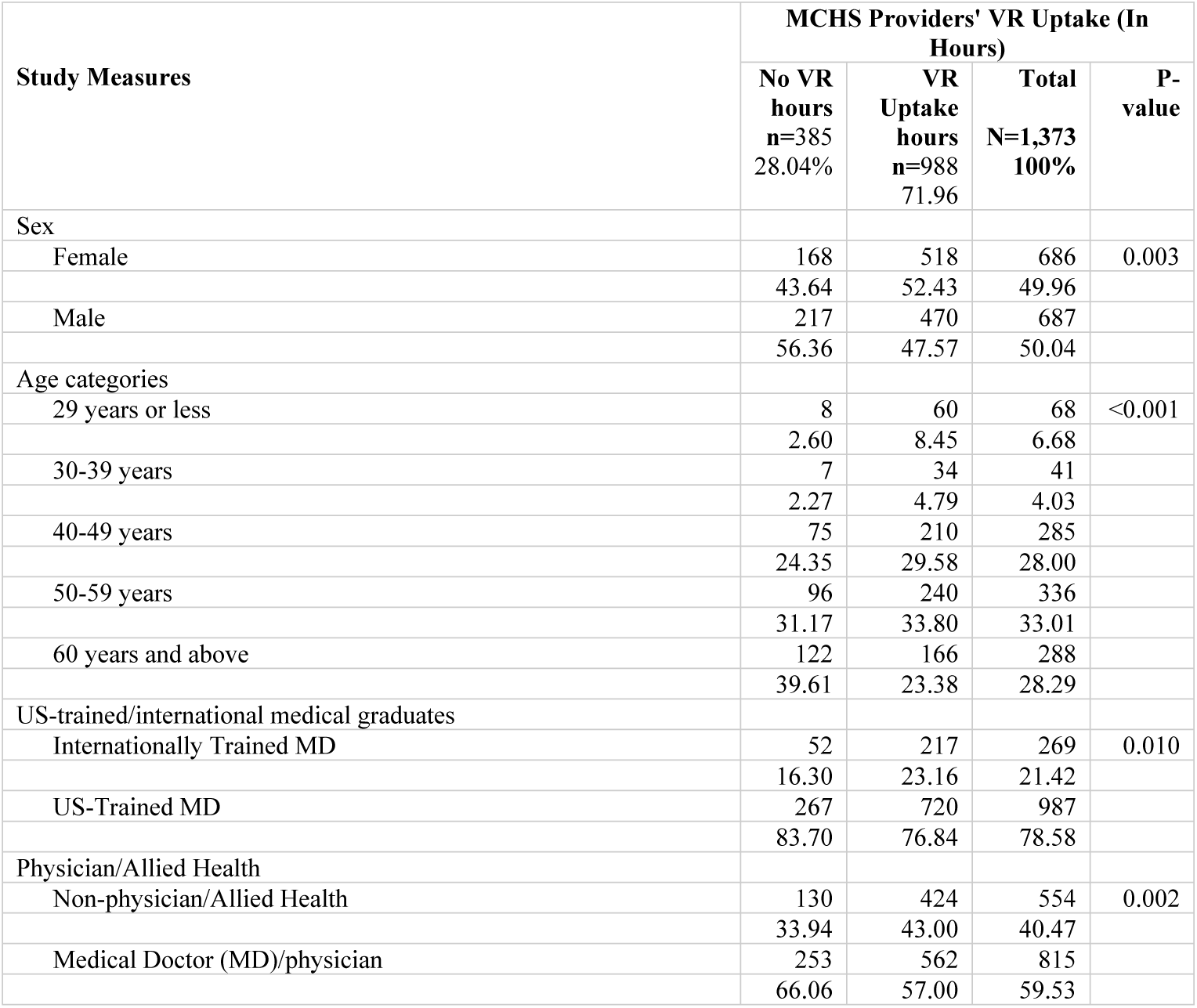
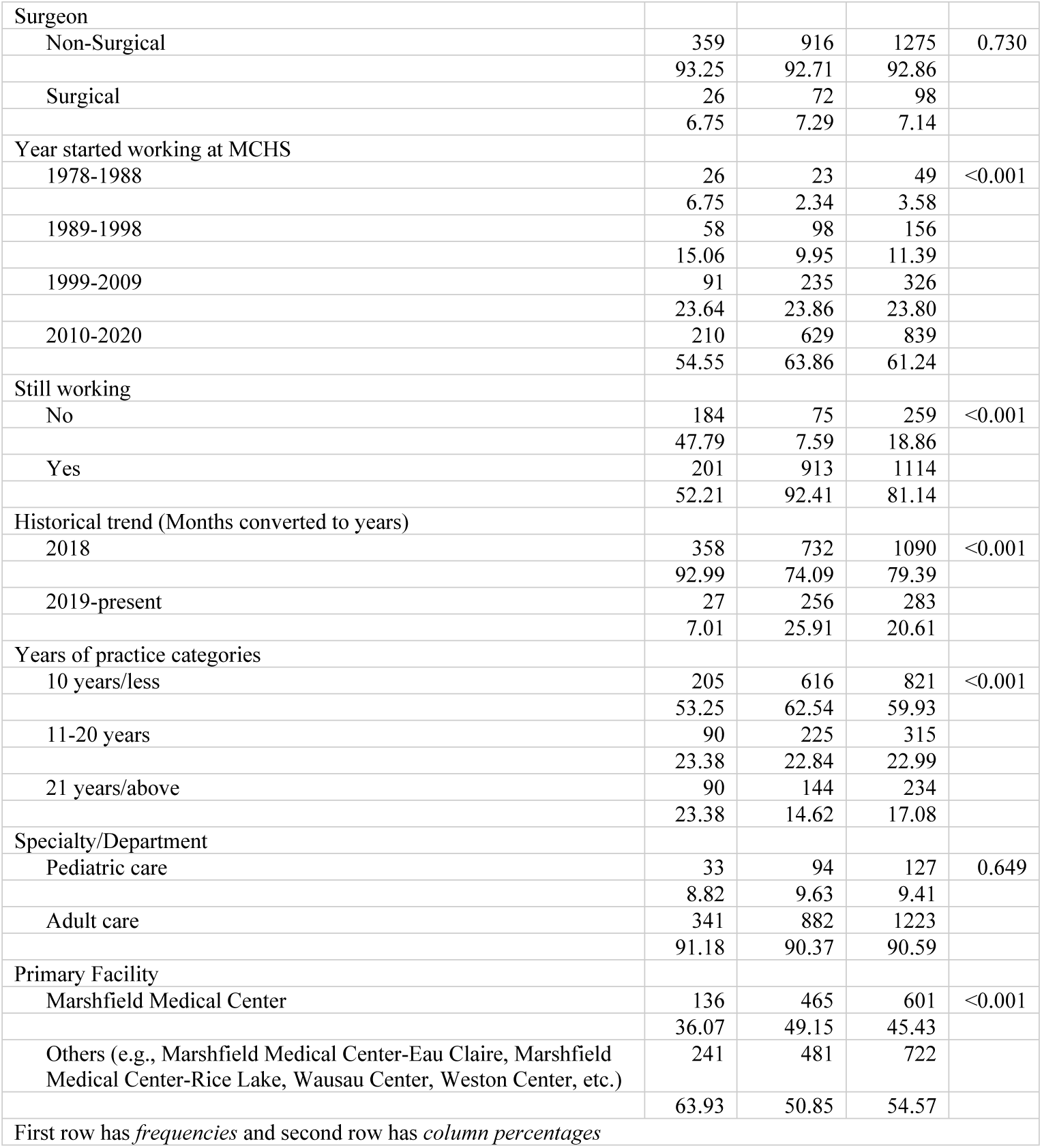
MCHS Providers’ Voice Recognition Uptake by User and Institutional Characteristics, 2018-2020.

Bivariate analysis indicates that physicians 60 years and older had the highest proportion of no voice recognition uptake (39.6%) of the 385 with no VR uptake, while those 29 years and younger had the lowest (2.6%), at p<0.001 (Fig 1). Bivariate analysis shows that US-trained physicians had the highest proportion of no voice recognition uptake (83.7%) of the 385 (28.0%) compared to internationally trained physicians (16.3%), at p=0.010 (Fig 2). Of the 385 MCHS providers with no voice recognition uptake, bivariate analysis indicates that females had a lower proportion (43.6%) compared to males (56.4%) providers, at p=0.003 (Fig 3).

**Fig 1:**
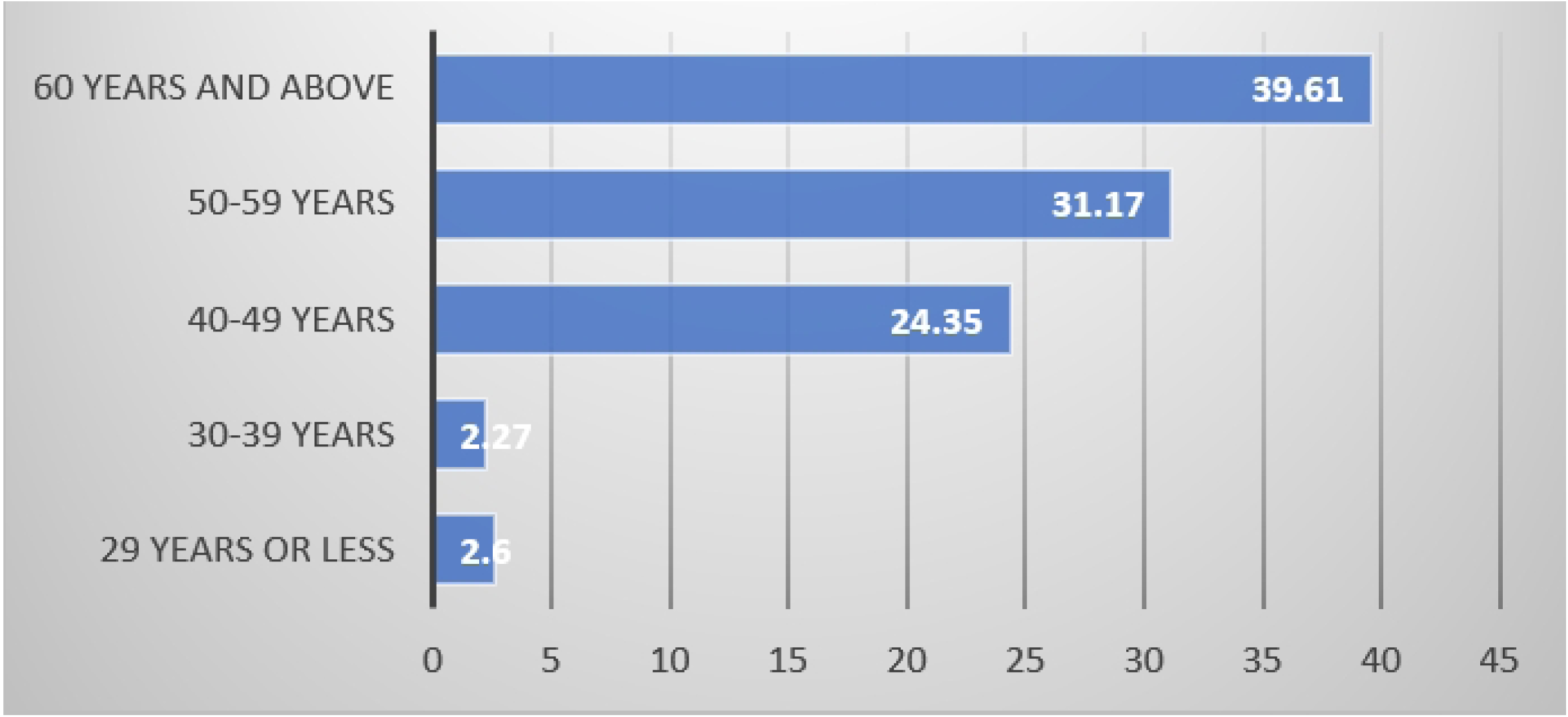
Percentage of No VR Uptake, by MCHS Provider Age (p<0.001) Bivariate analysis indicates that physicians 60 years and older had the highest proportion of no voice recognition uptake (39.6%) of the 385 (28.04%) providers with no VR uptake, while those 29 years and younger had the lowest (2.6%), at p<0.001.

**Fig 2:**
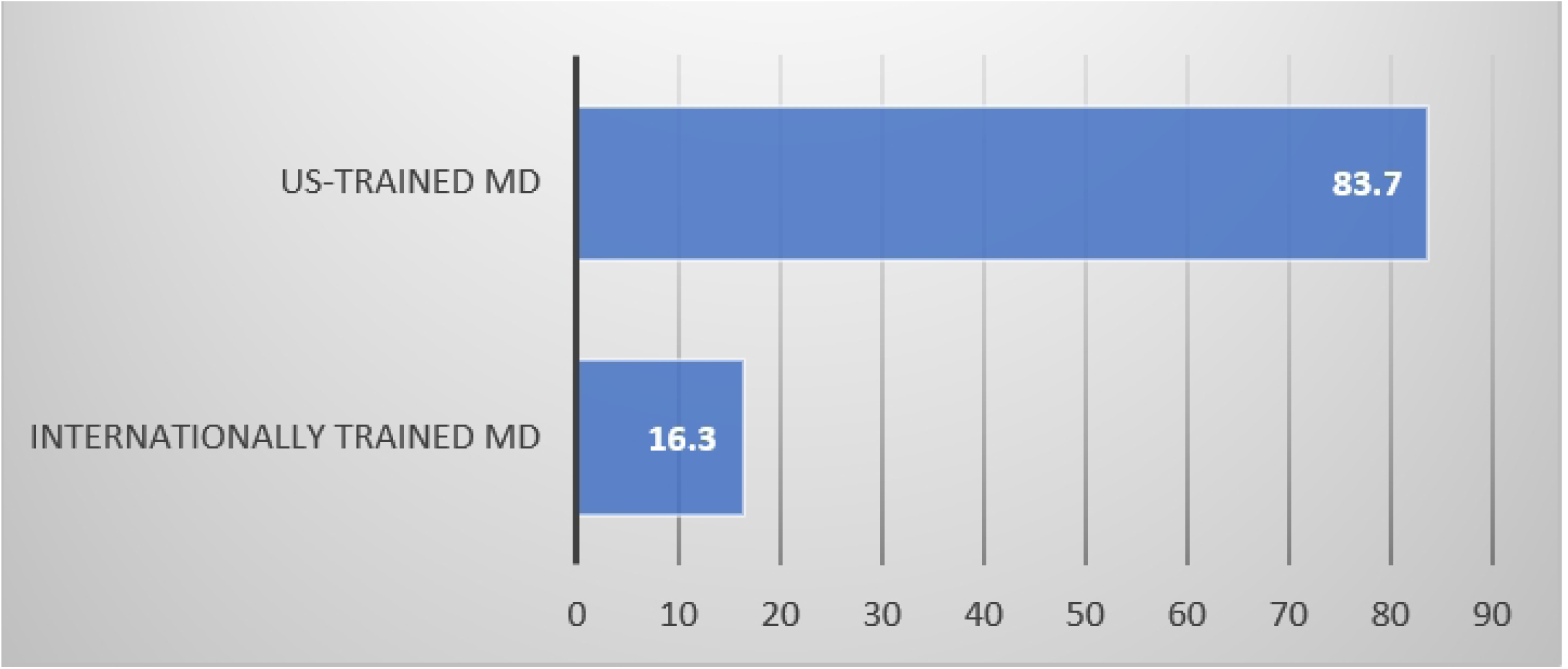
Percentage of No VR Uptake, by US-trained/intemational medical graduates (p=0.010) Bivariate analysis shows that US-trained physicians had the highest proportion of no voice recognition uptake (83.7%) of the 385 (28.04%) providers compared to internationally trained physicians (16.3%), at p=0.010.

**Fig 3:**
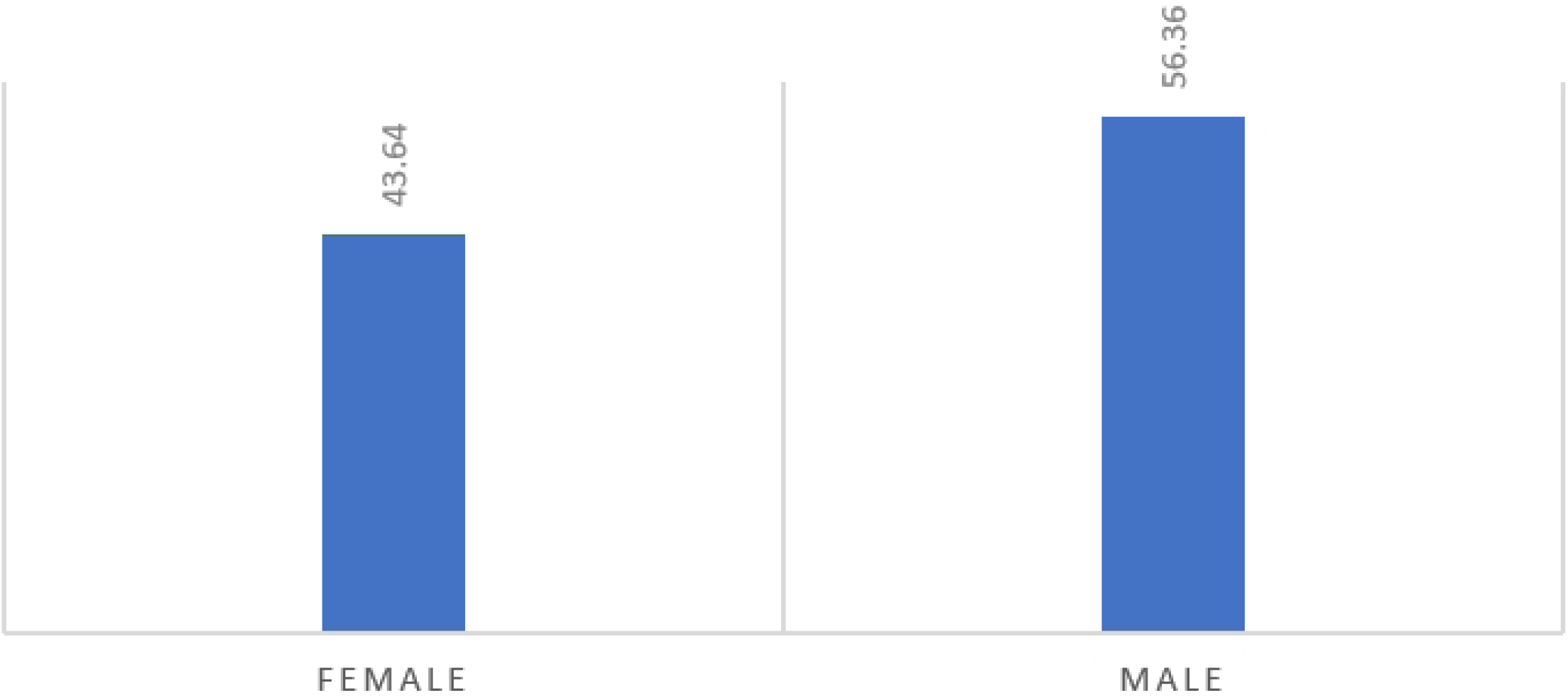
Percentage of No VR Uptake, By MCHS Provider Gender (P=0.003) Of the 385 (28.04%) providers with no voice recognition uptake, bivariate analysis indicates that female providers had a lower proportion (43.64%) compared to male (56.36%) providers, at p=0.003.

Table 3 presents a multivariate analysis of user and institutional factors influencing MCHS provider VR uptake (study outcome) for all users and facilities from 2018 to 2020. When compared to 2018, the odds of VR uptake were 4.9 times significantly higher in 2019 and 2020 (AOR: 4.99, 95% CI: 2.53 - 9.88). Physicians trained in the US had a 0.6 lower likelihood of VR uptake when compared to internationally trained physicians (AOR: 0.60, 95% CI: 0.36 - 0.99).

**Table 3:** User and Institutional Factors Influencing MCHS Provider Voice Recognition Uptake (Study Outcome), Adjusted, 2018-2020.

| Study Measures | OR | 95% CI |  | Sig |
| --- | --- | --- | --- | --- |
| Sex: Female | Ref. |  |  |  |
| Male | 1.081 | .737 | 1.587 |  |
| Age categories: 29 years/less | Ref. |  |  |  |
| 30-39 years | .883 | .217 | 3.602 |  |
| 40-49 years | .618 | .205 | 1.859 |  |
| 50-59 years | .557 | .183 | 1.694 |  |
| 60 years/above | .311 | .101 | .961 | ** |
| US-trained/international medical graduates: International MD | Ref. |  |  |  |
| US-Trained MD | .596 | .360 | .985 | ** |
| Physician/Allied Health: Medical Doctor (MD)/physician | Ref. |  |  |  |
| Non-physician/Allied Health | .734 | .481 | 1.120 |  |
| Year started working at MCHS: 1978-1988 | Ref. |  |  |  |
| 1989-1998 | 1.727 | .772 | 3.861 |  |
| 1999-2009 | 1.941 | .603 | 6.248 |  |
| 2010-2020 | .749 | .193 | 2.915 |  |
| Still working: No | Ref. |  |  |  |
| Yes | 15.952 | 9.792 | 25.987 | *** |
| Years of practice categories: 10 years/less | Ref. |  |  |  |
| 11-20 years | .381 | .137 | 1.061 | * |
| 21 years/above | .367 | .115 | 1.175 | * |
| Primary Facility: Marshfield Medical Center | Ref. |  |  |  |
| Others (e.g., Marshfield Medical Center-Eau Claire, Marshfield Medical Center-Rice Lake, | .479 | .334 | .686 | *** |
| Wausau Center, Weston Center, etc.) |  |  |  |  |
| Historical trend (Months converted to years): 2018<br>2019-present | Ref.<br>4.997 | 2.527 | 9.881 | *** |
| *** $p < .01$, ** $p < .05$, * $p < .1$ | | | | |

Table 4 presents an unadjusted and adjusted analysis of user and institutional factors influencing VR uptake among MCHS physicians only from 2018 to 2020. In both the unadjusted and adjusted analyses, the likelihood of VR uptake was 0.5 times (OR: 0.53, 95% CI: 0.37 - 0.76) and 0.6 times (AOR: 0.58, 95% CI: 0.35 - 0.97) lower among US-trained physicians compared to internationally trained physicians, respectively. VR uptake was significantly associated with other physician-related factors in both the unadjusted and adjusted analyses. The likelihood of VR uptake among physicians was 4.3 times higher in 2019 and 2020 compared to 2018 (OR: 4.30, 95% CI: 2.44 - 7.46) in the unadjusted analysis and 7.7 times higher in the adjusted analysis (AOR: 7.74, 95% CI: 2.51 - 23.86). The likelihood of VR uptake was 0.2 times (OR: 0.20, 95% CI: 0.10 - 0.59) lower in the unadjusted analysis and 0.2 times (AOR: 0.17, 95% CI:

**Table 4:** Unadjusted and Adjusted analyses of User and Institutional Factors Influencing VR Uptake (Study Outcome) among MCHS Physicians, 2018-2020.

| Study Measures | Unadjusted |  |  |  | Adjusted |  |  |  |
| --- | --- | --- | --- | --- | --- | --- | --- | --- |
|  | OR | 95% CI |  | Sig | OR | 95% CI |  | Sig |
| US-trained/international medical graduates:<br>International MD | Ref. |  |  |  | Ref. |  |  |  |
| US-Trained MD | .529 | .368 | .761 | *** | .581 | .347 | .972 | ** |
| Sex: Female | Ref. |  |  |  |  |  |  |  |
| Male | .786 | .571 | 1.081 |  | .984 | .602 | 1.61 |  |
| Age categories: 29 years/less | Ref. |  |  |  | Ref. |  |  |  |
| 30-39 years | .720 | .159 | 3.268 |  | .335 | .04 | 2.807 |  |
| 40-49 years | .373 | .123 | 1.135 | * | .330 | .054 | 2.003 |  |
| 50-59 years | .407 | .136 | 1.222 |  | .439 | .070 | 2.754 |  |
| 60 years/above | .197 | .066 | .585 | *** | .169 | .0270 | 1.063 | * |
| Surgical/non-surgical: Non-Surgical | Ref. |  |  |  |  |  |  |  |
| Surgical | 1.331 | .807 | 2.193 |  |  |  |  |  |
| Year started working at MCHS: 1978-1988 | Ref. |  |  |  | Ref |  |  |  |
| 1989-1998 | 3.117 | 1.431 | 6.789 | *** | 2.095 | .835 | 5.260 |  |
| 1999-2009 | 3.217 | 1.529 | 6.766 | *** | 2.176 | .400 | 11.837 |  |
| 2010-2020 | 5.119 | 2.515 | 10.418 | *** | 3.808 | .356 | 40.770 |  |
| Still working: No | Ref. |  |  |  | Ref |  |  |  |
| Yes | 9.578 | 6.569 | 13.964 | *** | 15.108 | 7.874 | 28.985 | *** |
| Historical trend (Months converted to years):<br>2018 | Ref. |  |  |  | Ref |  |  |  |
| 2019-present | 4.262 | 2.435 | 7.459 | *** | 7.743 | 2.512 | 23.864 | *** |
| Years of practice categories: 10 years/less | Ref. |  |  |  | Ref |  |  |  |
| 11-20 years | .646 | .445 | .938 | ** | 1.254 | .180 | 8.725 |  |
| 21 years/above | .514 | .354 | .746 | *** | 1.658 | .174 | 15.786 |  |
| Specialty/Department: Pediatric Care<br>Specialties | Ref. |  |  |  |  |  |  |  |
| Adult Care Specialties | .779 | .495 | 1.226 |  |  |  |  |  |
| Primary Facility: Marshfield Medical Center | Ref. |  |  |  | Ref |  |  |  |
| Others (e.g., Marshfield Medical Center-Eau Claire, Marshfield Medical Center-Rice Lake, Wausau Center, Weston Center,<br>etc.) | .463 | .341 | .630 | *** | .434 | .273 | .690 | *** |
| The variables in the adjusted model have blank spaces because they were not significant in the unadjusted analyses.<br>*** $p<.01$. ** $p<.05$. * $p<.1$ | | | | | | | | |

0.27 - 1.06) lower in the adjusted analysis among physicians aged 60 and over compared to physicians aged 29 and under, respectively. When compared to Marshfield Medical Center, the likelihood of VR uptake was 0.5 times (OR: 0.46, 95% CI: 0.34 - 0.63) lower in the unadjusted analysis and 0.4 times (AOR: 0.43, 95% CI: 0.27 - 0.69) lower in the adjusted analysis among Others (e.g., Marshfield Medical Center-Eau Claire, Marshfield Medical Center-Rice Lake, Wausau Center, Weston Center, etc.).

## Discussion

When physicians and allied health staff navigate charts and enter or retrieve data using conventional keyboard and mouse interfaces, usability issues in the EHR can result in workflow inefficiencies. Voice input technology allows another method of EHR and has been utilized to sidestep some problems with traditional interfaces (11). The current body of literature has focused on VR use in specific hospital units, such as the emergency department and radiology, while a few have looked at the impact of VR systems across healthcare systems (3,12,13). Furthermore, the majority of studies did not describe user characteristics (14–21). Due to user homogeneity, the studies that did analyze user characteristics such as age, gender, or native language lacked statistical power to examine the effect of user characteristics on VR adoption (13,22,23). Although some studies have described contributing factors for the various accuracy rates of VR systems, including adequate vocabularies for specific medical domains, speaking fluency, background noise of the clinic environment, and comparable instruments connecting to the RTD system (3), no systematic examination has been conducted regarding how individual and institute levels influence VR uptake of new documentation technologies. Our study fills this gap using a retrospective approach to examine VR uptake across an entire healthcare system and evaluate the VR uptake by individual/institute characteristics. Most studies did not examine the characteristics or only looked at a few, such as sex and specialty (3,22). Our study adds healthcare providers’ backgrounds (i.e., training, years of clinical working experience) alongside institute features (locations). This investigation reflects the significant reality of heterogeneous healthcare providers in rural areas.

Our results indicate that VR uptake across MCHS was significantly higher than no VR uptake among the 1,373 providers studied (72 percent vs. 28 percent). According to the adjusted analysis, VR adoption increased significantly across MCHS since 2018 despite the fact that it was voluntary. However, VR uptake was 0.2 times lower among physicians aged 60 and over compared to physicians aged 29 and younger. A recent study (23) discovered that 82% of physicians were initially optimistic about using speech recognition technology with electronic medical records. After six months of use, 87 percent of physicians thought speech recognition technology was a good idea. Furthermore, in this study, 72 percent of the physicians expected speech recognition technology to save them time, and 51 percent of participants reported time savings after using it in the clinical setting. Increased acceptance of speech recognition technology among physicians has been attributed to technological advancements and the electronic medical record (23). Another cross-sectional study of 3473 physicians discovered that more than 94% planned to incorporate virtual health care into their practice by December 2020, and physicians born between 1928 and 1945 (Silent Generation) were less likely to be early adopters of new virtual health technologies (24). Although age differences are unsurprising, the reluctance of any physician to adopt new technologies or to adopt them slowly risks workflow interruptions and communication problems that can affect patient care.

Foreign-trained physicians accounted for approximately 25% of the specialties actively serving patients in the United States, and immigration regulations actively recruit and retain them to practice in underserved areas (6,7). Foreign-trained physicians must pass the United States Medical Licensing Examination and complete residencies regardless of where they have trained. Until now, the use of VR technologies in assisting patient care documentation in health systems with second-language users has been understudied. Our adjusted model results indicate that the likelihood of VR adoption is 0.6 times lower among US-trained physicians than among internationally trained physicians, despite the fact that internationally trained physicians are in the minority in our study, accounting for only 21.4 percent of the MCHS workforce. Indeed, MCHS has a higher proportion of overseas-trained healthcare providers caring for our patients than other US health systems, but this finding shed light on more specific healthcare issues and conditions. Due to growing number of foreign-trained or international-trained physicians filling medical gaps in the US (6,25), this finding is a pioneer in studying VR technologies among a diverse set of providers. Utilizing VR technologies might be a challenge for foreign-trained or international-trained healthcare providers due to multiple adaptations they make navigating second-language acquisition. VR software/RTD systems were originally developed by native speakers; higher error rates were associated with second-language users (26). But according to our findings, VR uptake was lower among US-trained physicians compared to internationally trained physicians. This finding suggests that foreign-trained or international-trained medical graduates not only play important roles in filling medical gaps in disadvantaged communities, but they may have also overcome the multiple adaptation barriers caused by second-language, indicating VR technologies can be used by a diverse range of healthcare providers. In fact, patient outcomes including mortality are significantly better among physicians who graduated from medical schools outside the United States than among those who graduated from medical schools in the United States (27). Data on older Medicare patients admitted to US hospitals revealed that patients treated by international medical graduates had lower mortality than patients treated by US graduates, despite those patients treated by having slightly more chronic conditions (27). International graduates may outperform US graduates in terms of patient outcomes for a variety of reasons. According to the 2015 National Resident Matching Program, the current approach to allowing international medical graduates to practice in the United States may select for better physicians on average, and international graduates who are successful in the US matching process may represent some of their home country’s best physicians. The fact that international graduate students outperform US graduates on test scores in the United States renders some support to this hypothesis (28). Furthermore, many international graduates who are currently practicing in the United States likely underwent residency training twice, once in their home country and once in the United States, and such intensive and prolonged training may be another reason for their superior performance (27).

Our findings also show gender differences in VR non-adoption. Bivariate analysis reveals that female providers made up a lower proportion (43.6 percent) of the 385 MCHS providers in our sample who did not use voice recognition than male providers (56.4 percent). This implies that males outnumber females among providers who did not use VR and that more female providers use (but are not necessarily more likely to use) VR than male providers. In contrast, research published by the North American Chapter of the Association for Computational Linguistics shows that there are significant gender biases in VR, that it effectively recognizes white male voices, and that, for example, Google’s VR system is 13 percent more accurate for men than it is for women (29). While we acknowledge that gender differences in VR accuracy have dire implications in the real world, gender disparities in our study may exist as a result of how our databases were structured. The underlying reason could be that databases contain a lot of (white) male data and less data on female and minority voices, though our study did not take race/ethnicity and accuracy measures into account, nor did it confirm this hypothesis because it included a nearly equal number of female (49.96 percent) and male (50.04 percent) providers.

However, it is important to note that databases that do not and have mostly (white) male voices will not perform as well as those that do and have more diverse voices, such as female and other more diverse voices, such as internationally trained physicians. This would then be a clear case of social injustice, which should compel healthcare providers to address the issue. As VR systems become more prevalent and powerful, they will have a greater impact on our daily lives. By creating a world where everyone’s voices are clearly heard, healthcare providers may increase VR adoption across diverse healthcare workforces. This could be achieved through research and transparency, as well as reporting accuracy rates for women and diverse users, and then citing how well the VR system understands those demographics.

Future research opportunities should be acknowledged. First, this study only examined VR uptake and trends in MCHS including use, and institutional factors between 2018 and 2020 after the policy was implemented, yet some other essential features were not included for further investigation. For example, the error rates of RTD and associated costs with documentation were not analyzed. At least one study has found that a higher error rate is found in speech recognition than in traditional dictation (30). Second, we investigated the number of providers who actively use VR to dictate. We did not, however, determine how many lines/hours were dictated by providers (by site/group/specialty), or what the average line per hour of providers (by site/group/specialty) is, or who the top and bottom users are in terms of total lines, total hours, and lines per hour. Answering these questions will assist in tracking and monitoring trends, as well as determining where additional enablement efforts are required or what impact additional enablement activities have had on VR adoption (9). Third, VR user efficiency measures, such as Auto-text and Voice Commands, were only described for the entire MCHS system and were not analyzed at the bivariate and multivariate levels. This means that our findings described how effectively VR users use VR efficiency features across the entire MCHS system, rather than drilling down into specific sites, groups, and specialties. To address this limitation, future research on this topic should examine how efficiently providers use advanced features such as Voice commands and Auto-Text (by site/group/specialty), what percentage of dictation is generated by advanced features such as Auto-Text (by site/group/specialty), and which providers could benefit from more training or assistance in dictating with Auto-Text or voice commands. Fourth, VR dictation quality, such as signal-to-noise ratio, was evaluated for the entire MCHS system. However, correlation analyses were not performed to identify potential training opportunities or environmental factors influencing a physician’s ability to use VR effectively. In most cases, in-person observation and discussion are required to determine what is impeding or affecting the physician so that appropriate corrective actions can be taken. Our study provides a starting point for determining the MCHS system average; however, viewing the information at an individual level allows us to target specific issues and determine how to improve the individual, group, site, and specialty levels appropriately. Future research is needed to investigate how background noise affects providers’ dictation quality, whether the user needs a refresher course on speech recognition best practices (e.g., speaking all punctuation), factors influencing Signal-to-Noise Ratio, and whether additional in-person observation is required to pinpoint the root cause of issues.

Our findings should assist providers and policymakers in identifying and developing targeted interventions to improve the VR adoption process across the system, as well as identifying the challenge of real-world VR application in other healthcare systems by exploring and gauging variations in VR uptake at the individual and institute/unit levels. This knowledge could eventually lead to better documentation, unit communication, and patient outcomes in healthcare systems. Finally, the study findings support the need to allocate additional resources for VR adoption to ensure high-quality care documentation, reduce time for patient information sharing processes, and improve patient care.

## Conclusion

Accurate clinical documentation is crucial to health care quality and safety. Clinicians in the United States rely heavily on dictation services supported by speech recognition technology and professional medical transcriptionists (12). Using data from the Nuance Management Center for 1,373 MCHS providers from 2018 to 2020, we assessed the uptake of VR for RTD in a rural healthcare system. VR adoption has increased significantly across MCHS and among physicians since 2018. However, VR adoption was lower among US-trained physicians than among international-trained physicians. VR uptake was also lower among physicians aged 60 and over compared to physicians aged 29 and less. These findings provide critical information about VR trends, physician factors, and which providers may benefit from additional training in order to increase VR adoption in healthcare systems in the United States.

## Data Availability

All relevant data are within the manuscript and its Supporting Information files.

## Abbreviations

VR: Voice Recognition
RTD: Real-Time Dictation
DMO: Dragon Medical One
NMC: Nuance Management Center
MCHS: Marshfield Clinic Health System
SD: Standard Deviation
OR: Odds Ratios
AOR: Adjusted Odds Ratios
CIs: Confidence Intervals

## Acknowledgments

We would like to thank the Marshfield Clinic Institutional Review Board for providing study ethical clearance to conduct this study. Our gratitude also goes to all data collectors and analysts who prepared the dataset for analysis, including Roxy Eibergen, Project Manager of Center for Clinical Epidemiology and Population Health.

## Supporting information

S1: Dataset (Available upon request)

## Notes

**Data Availability Statement:** All relevant data are within the manuscript and its Supporting Information files.

### Competing Interest Statement

The authors have declared no competing interest.

### Clinical Trial

Does not apply

### Funding Statement

The author(s) received no specific funding for this work.

### Author Declarations

This study was approved by Marshfield Clinic Institutional Review Board (IRB-20-718).

